# Waist-to-height ratio is a more accurate screening tool for hypertension than waist-to-hip circumference and BMI in type 2 diabetes: A prospective study

**DOI:** 10.1101/2020.09.29.20203752

**Authors:** Fatemeh Moosaie, Seyede Marzie Fatemi Abhari, Niloofar Deravi, Arman Karimi Behnagh, Sadaf Esteghamati, Fatemeh Dehghani Firouzabadi, Soghra Rabizadeh, Manouchehr Nakhjavani, Alireza Esteghamati

**Author notes:** Corresponding author: Alireza Esteghamati, MD, Professor of Endocrinology, Endocrinology and Metabolism Research Center (EMRC), Vali-Asr Hospital, Tehran University of Medical Sciences, Tehran, Iran, P.O. Box: 13145-784.

## Abstract

**Background:** To date, anthropometric measures (i.e. body mass index (BMI), waist to hip ratio (WHR) and waist to height ratio (WHtR) have shown to be associated with prediction of incident hypertension. However, the difference in accuracy of these measures has been of controversy. We aimed to determine whether WHtR is a more accurate tool for HTN than WHR and BMI in patients with type 2 diabetes.

**Material and Methods:** The study population consisted of 1685 normotensive patients with T2DM. They were followed up for hypertension incidence for a mean of 4.8 years from April 2002 to January 2020. Cox regression was performed to assess the association between anthropometric measures (i.e., BMI, WHR, and WHtR) and incident hypertension during the follow-up period. Area under the ROC curve analysis was performed and optimal cutoff values were calculated using Youden index.

**Results:** WHtR and BMI were significantly associated with an increased risk of hypertension (HR=3.296(0.936-12.857), P < 0.001 and HR:1.050 (1.030-1.070), P < 0.001, respectively). The discriminative powers of each anthropometric index for HTN were 0.571 (0.540–0.602) for BMI, 0.518 (0.486–0.550) for WHR, and 0.609 (0.578–0.639) for WHtR. The optimal cutoff points for predicting HTN in patients with T2DM were 26.94 (sensitivity=0.739, specificity=0.380) for BMI, 0.90 (sensitivity=0.718, specificity=0.279) for WHR and 0.59 (sensitivity=0.676, specificity=0.517) for WHtR.

**Conclusion:** In the current study WHtR was a more accurate screening tool for HTN compared to WHR and BMI in patients with type 2 diabetes.

## Introduction

Diabetes mellitus is among the most prevalent diseases worldwide, which significantly affects the cardiovascular system. The main cause of death among patients with diabetes is reported to be directly related to cardiovascular diseases (CVD) [1-3]. Diabetes mellitus is associated with several major cardiovascular complications, including coronary artery disease, stroke, ischemic heart disease, myocardial infarction, and heart failure. Cardiovascular risk factors increase the risk of occurrence of these complications. Hypertension (HTN), a highly prevalent cardiovascular risk factor in patients with diabetes (present in 35% of men and 46% of women with diabetes), causes three forth of the cardiovascular deaths in these patients [4]. The hypertension diagnosis is confirmed in a gradual process requiring multiple careful blood pressure assessments at different times [5]. Late hypertension diagnosis extends the high blood pressure harmful effects on the cardiovascular system, promoting the onset of irrecoverable sequelae [6].

Since obesity plays a significant role in the etiology of hypertension, mainly in the abdominal obesity, using anthropometric indicators of adiposity may help screen patients who are at a higher risk for hypertension. Thus, the identified individuals may be referred earlier to health centers with better structures for hypertension monitoring and early diagnosis in order to optimize the opportunity for secondary prevention actions [7].

Body mass index (BMI), the standard general obesity measure, is the most commonly used indicator in hypertension prediction and screening. Adipose tissue distribution is more vital in the cardiovascular diseases compared with total body fat; thus, many other measures, including WHR (waist-to-hip ratio) and WHtR (waist-to-height ratio), have been developed which consider the distribution of adipose tissue [8]. Researchers have conducted many studies to assess the associations between several obesity indicators and hypertension in recent decades. Cuban and Japanese studies suggested BMI as the best single indicator of hypertension [9, 10]. In contrast, an Iranian cross-sectional study suggested WHR combined with BMI as the best predictor of hypertension in women [11]. A meta-analysis in 2006 reported the superiority of centralized obesity measures, especially WHtR over BMI for CVD risk detection [12]; however, another meta-analysis in 2008 reported that none of the anthropometric variables were systematically better than the others at the hypertension discrimination [13]. Therefore, it has remained controversial, which measure is the best predictor of hypertension. The present prospective study aimed to determine whether WHtR is a better screening tool for HTN than WHR and BMI in patients with type 2 diabetes.

## Methods

### Participants

Four thousand fifty-five (4055) patients who were previously diagnosed with type 2 diabetes voluntarily participated in this study from April 2002 to January 2020. All patients attended the endocrinology clinic of Vali-Asr Hospital, a tertiary center affiliated with Tehran University of Medical Sciences. After excluding patients with previously diagnosed hypertension (N= 2362), 1693 patients remained. Of these patients, 8 were excluded due to either failure to attend monthly follow-up or missing data on either systolic blood pressure, weight, height, or waist circumference. The final study population consisted of 1685 normotensive patients with T2DM (**Figure 1**). The patients’ anthropometric measures, including BMI, WHR, and WHtR, were assessed at baseline. They were followed up for hypertension incidence for a mean follow up period of 4.8 years from April 2002 to January 2020. This study was in full compliance with the Declaration of Helsinki and was reviewed and approved by the ethics committee of Tehran University of Medical Sciences.

**Figure 1.**
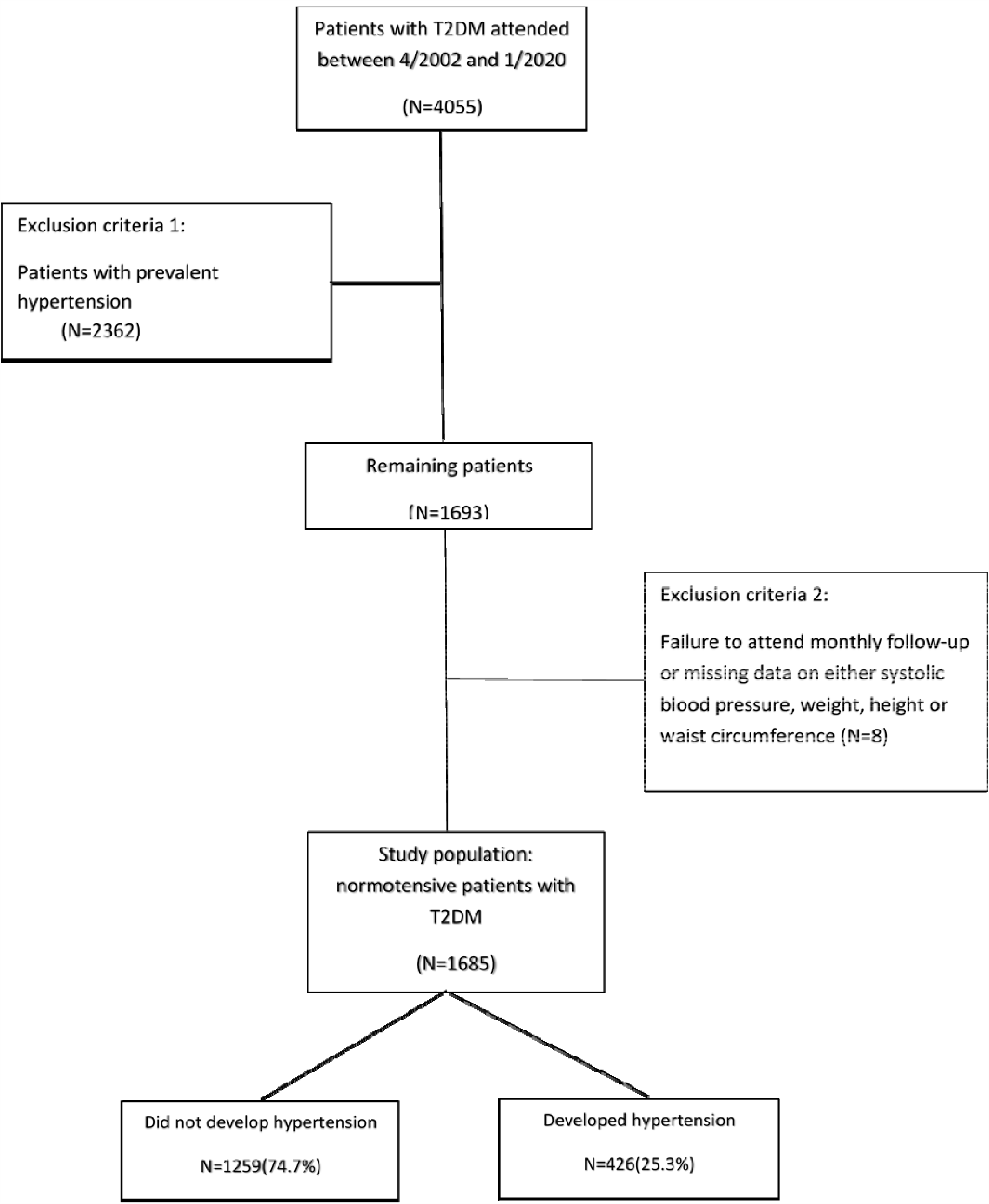
Study population selection flow chart; T2DM= type 2 diabetes mellitus

### Data Collection

At baseline, all participants were interviewed by a trained interviewer. Data collection at baseline consisted of the following parameters: identification and sociodemographic characteristics including age, gender, education, marital status; lifestyle habits such as smoking; self-reported health conditions and specifically the use of antihypertensive drugs; past medical history including any physical or mental conditions; and objective measures including blood pressure and body weight. Data collection during the follow-up period was limited to the following parameters: identification (only age and gender), self-reported health conditions (use of antihypertensive medications), and objective measures (only blood pressure). In this study, current smokers were defined as individuals who had smoked a minimum of 100 cigarettes in their lifetime and continued smoking during the follow-up period. A team of trained and closely observed researchers using standard equipment was responsible for data collection at all times.

### Anthropometric Measurements

Weight was measured using a portable digital scale with a precision of 0.1 kg in minimally clothed participants without shoes. Height was measured using a standard protocol with a tape measure. The body mass index (BMI) was calculated as weight (kilograms) divided by squared height (m^2^). Based on WHO classification the BMI values were categorized as follow: < 18.5 kg/m2 (underweight), 18.5 to 24.9 kg/m2 (normal weight), 25 to 29.9 kg/m2 (overweight) and ≥ 30 kg/m2 (obese). Waist circumference (WC) was measured at the end of a normal expiration. The tape was positioned horizontally at the midway between the iliac crest and the inferior border of the last rib. Likewise, Hip circumference was measured at the widest circumference of buttock while the tape was horizontal and parallel to the ground. During these procedures, the subjects were standing and wearing minimal clothing. All the measurements were performed with a precision of 0.1 cm. In order to minimize observer error, all measurements were carried out by the same technician. Waist to height (WHtR) and waist to hip ratio (WHR) were calculated by dividing waist (cm) by height (cm) and hip circumference (cm), respectively.

### Laboratory Measurements

From each subject, 10mL of venous blood was drawn after 12-14 hours of overnight fasting. The samples were kept in cold biochemistry tubes with a temperature of 4–8°C and sent to the representative calibrating laboratories within 4 hours, where the samples were immediately centrifuged (1500 Rmp, for 10 mins at standard room temperature 21°C). Then, the extracted serum was used for laboratory evaluations. Enzymatic calorimetric methods using the glucose oxidase test were implemented to measure the fasting plasma glucose (FPG) and 2h post-prandial glucose (2hPPG). Glycated hemoglobin (HbA1c) was measured using high-performance liquid chromatography. By using enzymatic methods, the serum concentrations of triglycerides, low-density lipoprotein cholesterol (LDL-C), high-density lipoprotein cholesterol (HDL-C), and total cholesterols were determined. The commercially-available kits, distributed by the central reference laboratory (Tehran, Iran), have passed all the quality control procedures.

### Definition of Hypertension

Blood pressure (BP) was measured on the left arm by using a mercury sphygmomanometer (Reishter, Germany) with appropriate cuff size according to the subjects’ arm circumference. The procedure was performed in the sitting position after a 10-min rest period, and all the subjects were asked not to smoke or consume caffeine 30 mins preceding the procedure commencement. BP was measured twice, with a 10-min interval between the two measurements. During this interval, the patients were asked to remain seated with their left hand placed at the heart level and their palm in the upward-facing position. The mean of the two measured values were considered as the patient’s BP. Auscultated systolic blood pressure (SBP) was defined as the first Korotkoff sound heard during cuff deflation, while diastolic blood pressure (DBP) was marked by the disappearance of the last Korotkoff sound. Patients using antihypertensive drugs or subjects with an SBP higher than 140mmHg or a DBP higher than 90mmHg were classified as hypertensive [14].

### Statistical Analysis

SPSS version 24.0 (SPSS Inc., Chicago, IL, USA) was employed for statistical analysis. We used Kolmogorov–Smirnov and Shapiro-Wilk normality tests, P-P plot, and histogram to test the normality of our study population. The null hypothesis was rejected for all the variables; thus, they were normal. To assess the association between different variables and incident hypertension, uni-variable analysis of potential continuous and categorical risk factors was performed using t-test and chi-square, respectively. Data are reported as mean ± standard deviation (SD) for continuous variables and as proportions for categorical variables. Cox regression analysis was performed to assess the association between anthropometric measures (i.e., BMI, WHR, and WHtR) and incident hypertension during the follow-up period. Hazard ratios (HR) were adjusted for the confounding variables (i.e., age, gender, history of coronary artery disease (CAD), SBP, DBP, FPG, family history of hypertension). The Cox regression analysis was also used to calculate HRs of hypertension and their 95% CIs with reference to a normal BMI and the first quartile of WHR and WHtR. The adjustment was done for age, gender, history of CAD, SBP, DBP, FPG, history of CAD and family history of hypertension. The discriminative powers of BMI, WHR, and WHtR were assessed by the area under the ROC curve analysis in the group with incident hypertension. The area under the ROC curve of the baseline BMI, WHR, and WHtR, and their 95% confidence intervals (CIs) were reported. Optimal cutoff values were calculated for each aforementioned anthropometric measure for the diagnosis of hypertension. Youden index was employed for calculating the optimal cutoff value. The sensitivity and specificity of each of the anthropometric measures were determined based on the calculated cutoff value. The risk of incident hypertension was compared among the BMI categories (i.e., underweight, normal, overweight, and obese) and the quartiles of WHR and WHtR. A two-sided p-value < 0.05 was considered necessary to reject the null hypothesis.

## Results

### Baseline Characteristics

A total of 1685 participants were enrolled in this study. The mean follow-up period was 4.8 years, and throughout this period, 426 participants (25.3%) developed hypertension. Table 1 summarizes the baseline characteristics of all participants based on the development of HTN. A comparison of study subgroups at baseline revealed higher means for age, SBP, DBP, 2-hpp, CAD prevalence, and anthropometric measures, as well as a higher frequency of HTN in family members of the subjects who developed HTN. Furthermore, the two groups differed significantly in terms of BMI and WHtR. The frequency of obese subjects was significantly higher in HTN group (42.8% vs. 37%), and also a higher proportion of patients who developed HTN were in the fourth quartiles of WHtR (37.6% vs. 24.9%) (**Table 1**).

**Table 1.**
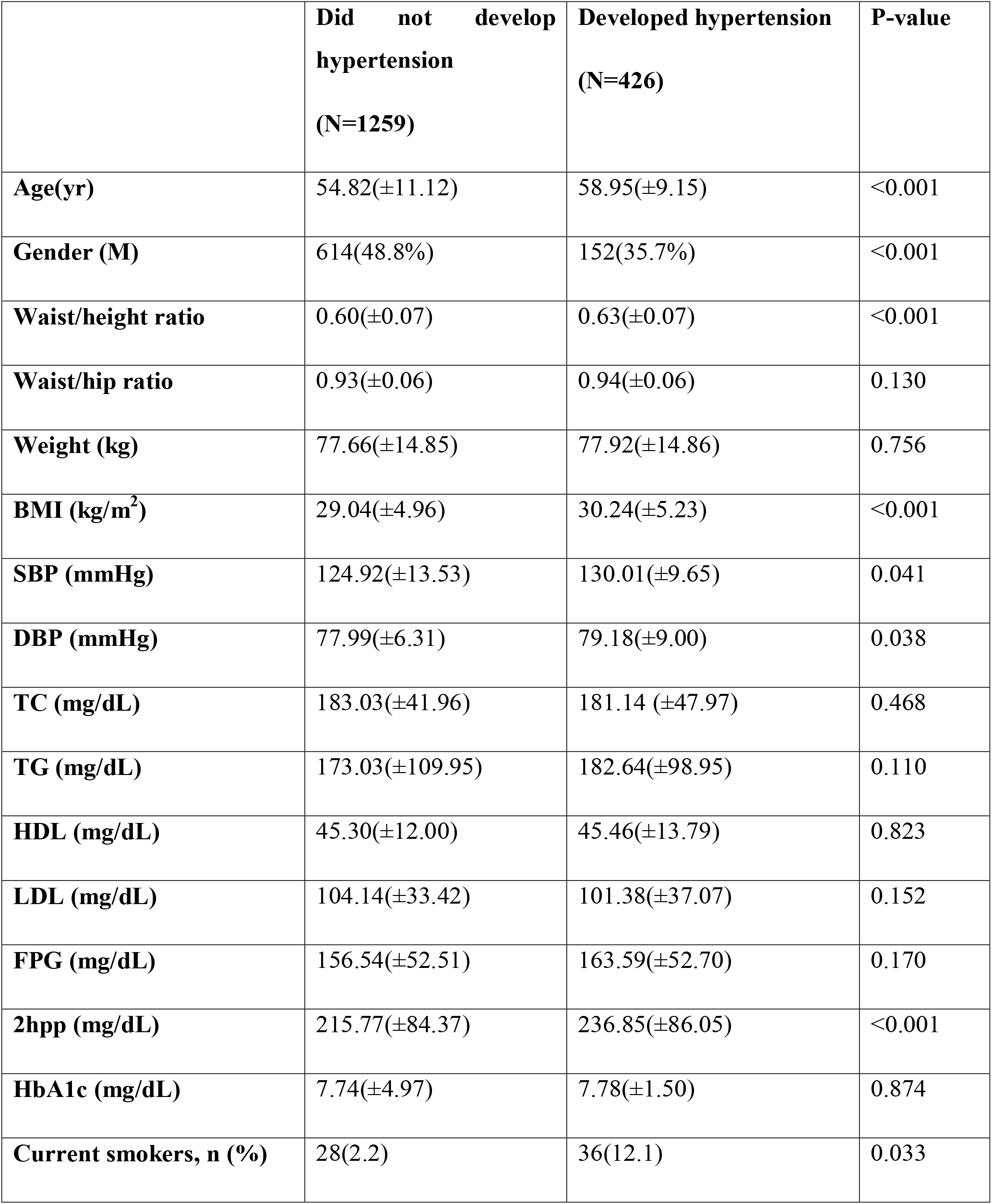
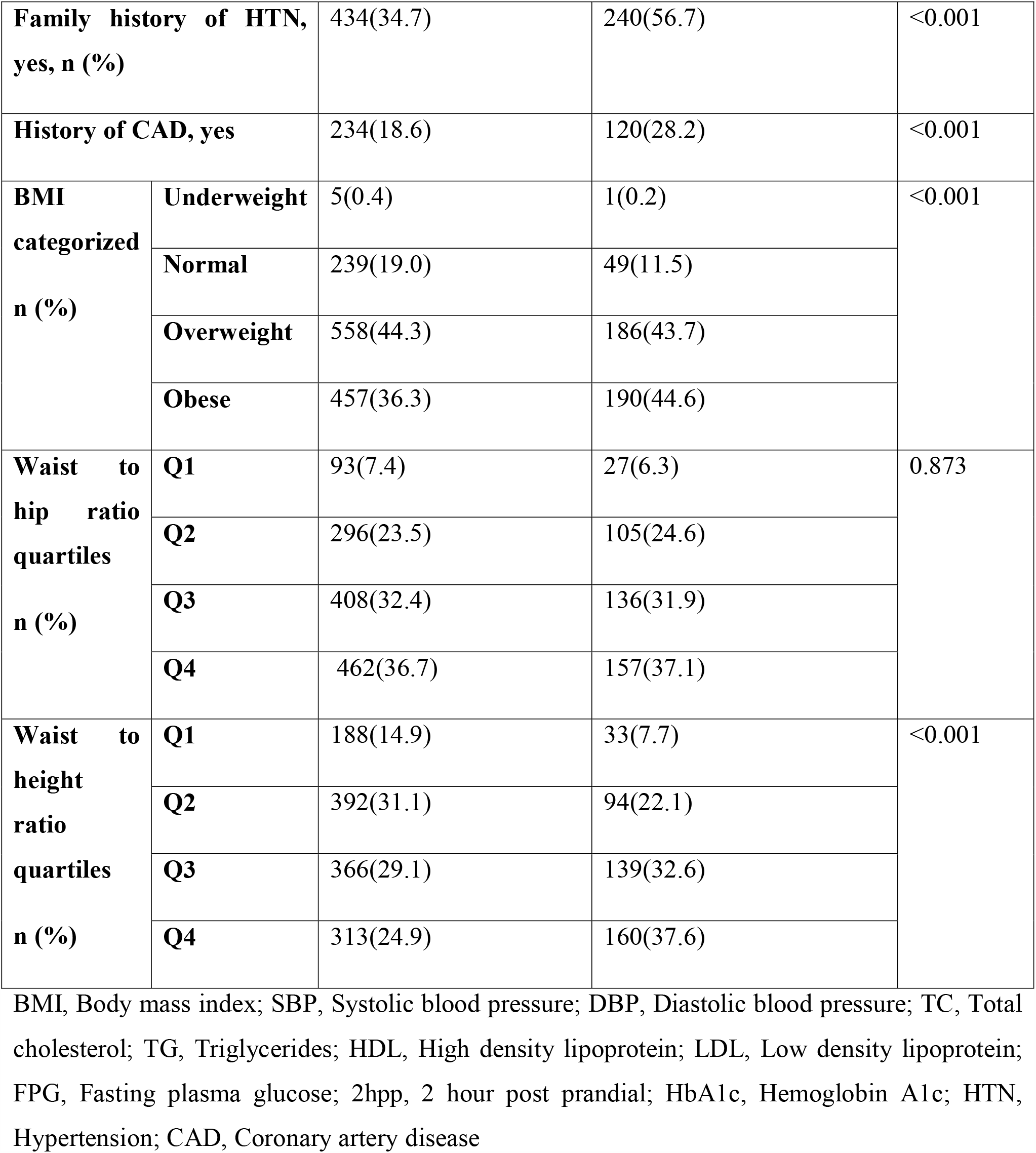
Baseline characteristics of the study populations, based on hypertension status at the time of follow-up.

|  | **Did not develop hypertension**<br>**(N=1259)** | **Developed hypertension**<br>**(N=426)** | **P-value** |
| --- | --- | --- | --- |
| **Age(yr)** | 54.82(±11.12) | 58.95(±9.15) | <0.001 |
| **Gender (M)** | 614(48.8%) | 152(35.7%) | <0.001 |
| **Waist/height ratio** | 0.60(±0.07) | 0.63(±0.07) | <0.001 |
| **Waist/hip ratio** | 0.93(±0.06) | 0.94(±0.06) | 0.130 |
| **Weight (kg)** | 77.66(±14.85) | 77.92(±14.86) | 0.756 |
| **BMI (kg/m<sup>2</sup>)** | 29.04(±4.96) | 30.24(±5.23) | <0.001 |
| **SBP (mmHg)** | 124.92(±13.53) | 130.01(±9.65) | 0.041 |
| **DBP (mmHg)** | 77.99(±6.31) | 79.18(±9.00) | 0.038 |
| **TC (mg/dL)** | 183.03(±41.96) | 181.14 (±47.97) | 0.468 |
| **TG (mg/dL)** | 173.03(±109.95) | 182.64(±98.95) | 0.110 |
| **HDL (mg/dL)** | 45.30(±12.00) | 45.46(±13.79) | 0.823 |
| **LDL (mg/dL)** | 104.14(±33.42) | 101.38(±37.07) | 0.152 |
| **FPG (mg/dL)** | 156.54(±52.51) | 163.59(±52.70) | 0.170 |
| **2hpp (mg/dL)** | 215.77(±84.37) | 236.85(±86.05) | <0.001 |
| **HbA1c (mg/dL)** | 7.74(±4.97) | 7.78(±1.50) | 0.874 |
| **Current smokers, n (%)** | 28(2.2) | 36(12.1) | 0.033 |

|  |  |  |  |  |
| --- | --- | --- | --- | --- |
| **Family history of HTN, yes, n (%)** |  | 434(34.7) | 240(56.7) | <0.001 |
| **History of CAD, yes** |  | 234(18.6) | 120(28.2) | <0.001 |
| **BMI categorized n (%)** | **Underweight** | 5(0.4) | 1(0.2) | <0.001 |
|  | **Normal** | 239(19.0) | 49(11.5) |  |
|  | **Overweight** | 558(44.3) | 186(43.7) |  |
|  | **Obese** | 457(36.3) | 190(44.6) |  |
| **Waist to hip ratio quartiles n (%)** | **Q1** | 93(7.4) | 27(6.3) | 0.873 |
|  | **Q2** | 296(23.5) | 105(24.6) |  |
|  | **Q3** | 408(32.4) | 136(31.9) |  |
|  | **Q4** | 462(36.7) | 157(37.1) |  |
| **Waist to height ratio quartiles n (%)** | **Q1** | 188(14.9) | 33(7.7) | <0.001 |
|  | **Q2** | 392(31.1) | 94(22.1) |  |
|  | **Q3** | 366(29.1) | 139(32.6) |  |
|  | **Q4** | 313(24.9) | 160(37.6) |  |

BMI, Body mass index; SBP, Systolic blood pressure; DBP, Diastolic blood pressure; TC, Total cholesterol; TG, Triglycerides; HDL, High density lipoprotein; LDL, Low density lipoprotein; FPG, Fasting plasma glucose; 2hpp, 2 hour post prandial; HbA1c, Hemoglobin A1c; HTN, Hypertension; CAD, Coronary artery disease

### Development of Hypertension in Patients with T2DM According to WHtR, WHR, and BMI

After adjusting for age, gender, history of CAD, SBP, DBP, FPG, and family history of HTN, WHtR and BMI were found to be significantly associated with an increased risk of hypertension (HR=3.296, 95%CI: 0.936-12.857, P < 0.001 and HR:1.050, 95%CI: 1.030-1.070, P < 0.001, respectively). However, this association was not significant for WHR (**Table 2**). In another adjusted cox regression model assessing the association between incident hypertension and different categories of BMI (i.e., underweight, normal, overweight, and obese), and four quartiles of WHR and WHtR, a higher WHtR at the baseline, was positively and significantly associated with the development of hypertension in a ratio-dependent manner (highest vs. lowest quartile HR 1.936, 95%CI: 1.306-2.871) (**Table 3**).

**Table 2.** Cox regression analysis determining the association between BMI, waist to hip ratio, and waist to height ratio with incident hypertension during the follow-up period; results are adjusted for age, gender, history of CAD, SBP, DBP, FPG and family history of HTN.

|  | **HR (95% CI)** | **P-value** |
| --- | --- | --- |
| **BMI** | 1.050(1.030-1.070) | <0.001 |
| **Waist to hip ratio** | 2.604(0.605-10.477) | 0.159 |
| **Waist to height ratio** | 3.296(0.936-12.857) | <0.001 |

BMI, Body mass index

**Table 3.** Hazard ratio of incident hypertension based on waist to height ratio, waist to hip ratio and BMI during the follow-up period; Data are presented as HR (95% CI); Results are adjusted for age, gender, history of CAD, SBP, DBP, FPG and family history of HTN.

|  | **Quartile 1** | **Quartile 2** | **Quartile 3** | **Quartile 4** | **P-value** |
| --- | --- | --- | --- | --- | --- |
| **Waist to hip ratio** | Ref | 1.247(0.812-1.914) | 1.039(0.680-1.587) | 1.246(0.810-1.917) | 0.328 |
| **Waist to height ratio** | Ref | 1.354(0.905-2.025) | 1.534(1.034-2.276) | 1.936(1.306-2.871) | 0.003 |
|  | **Underweight** | **Normal** | **Overweight** | **Obese** | **P-value** |
| **BMI** | 0.664(0.091-4.867) | Ref | 1.394(1.013-1.919) | 1.793(1.294-2.486) | 0.002 |

BMI, Body mass index

### ROC Analysis of WHtR, WHR, and BMI for Prediction of Hypertension in Patients with T2DM

**Figure 2** shows the areas under the ROC curve (AUC) of the anthropometric measurements for the prediction of HTN. All anthropometric indices, especially WHtR, exhibited a predictive power in detecting the incidence of HTN in patients with T2DM. The discriminative powers of each anthropometric index for HTN were 0.571 (95% CI: 0.540–0.602) for BMI, 0.518 (95% CI: 0.486–0.550) for WHR, and 0.609 (95% CI: 0.578–0.639) for WHtR. The optimal cutoff points for predicting HTN in patients with T2DM were 26.94 (sensitivity=0.739, specificity=0.380) for BMI, 0.90 (sensitivity= 0.718, specificity=0.279) for WHR and 0.59 (sensitivity=0.676, specificity=0.517) for WHtR.

**Figure 2.**
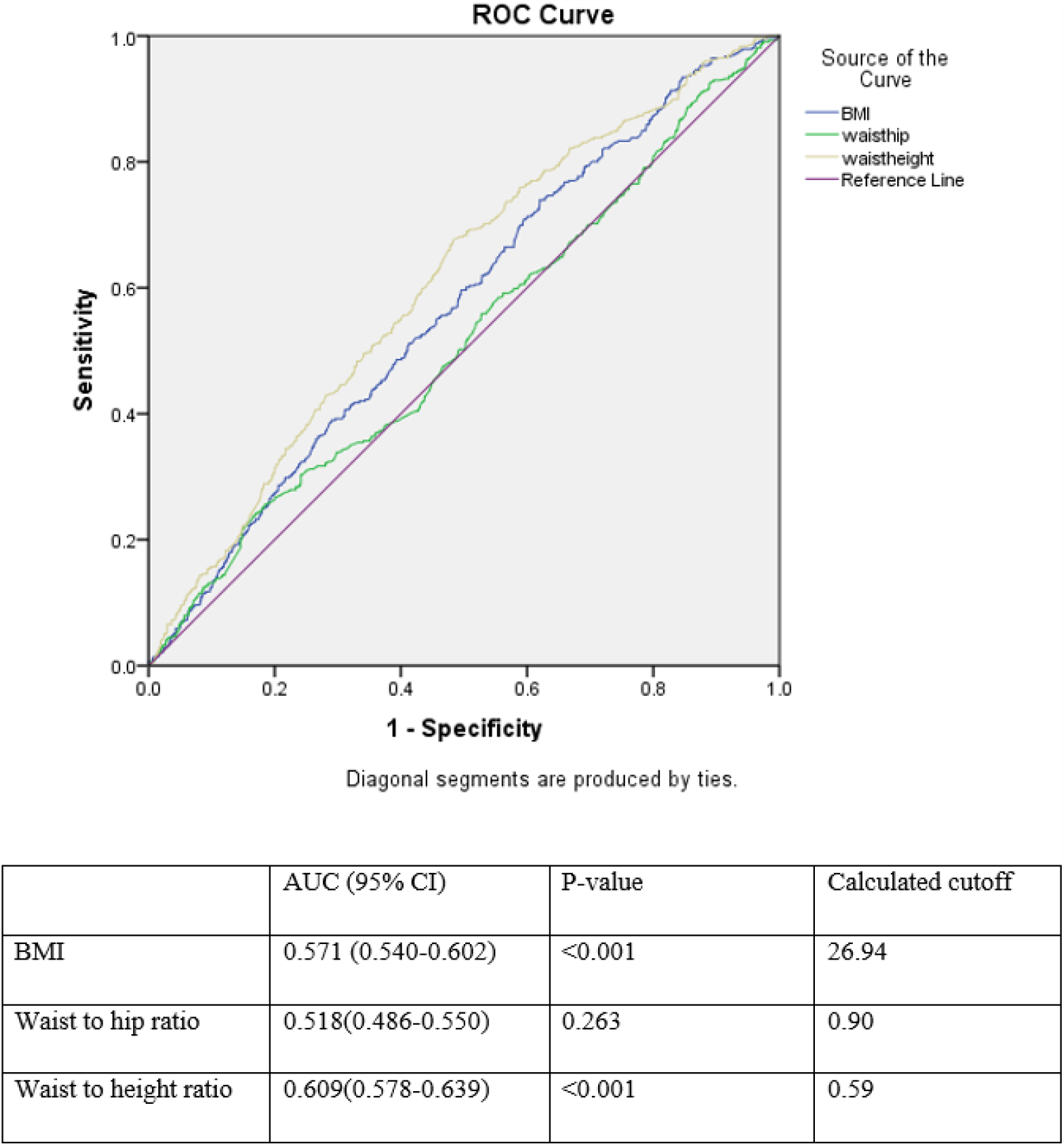
Receiver operating characteristics curve analysis for comparison of BMI, waist to hip ratio, and waist to height ratio as an index for incident hypertension

## Discussion

More than 4.5 million Iranian adults were diagnosed with diabetes in 2011; this number is projected to rise to 9.2 million by 2030. This increase emphasizes the significance of type 2 diabetes, most importantly, when considering the burden of diabetes-related complications [15]. Several studies have reported a rise in the record of hypertension in patients with diabetes. The existence of hypertension in patients with diabetes significantly increases the risk of stroke, coronary heart disease, retinopathy, and nephropathy [16]. Therefore, identifying risk factors and predictors of hypertension is of high importance to diagnose, prevent, control, and treat this condition as soon as possible. Our results indicated higher means for age, TG, CAD prevalence, smoking status and anthropometric measures, as well as a higher frequency of HTN in family members of the subjects who developed HTN. Additionally, the frequency of obese subjects was significantly higher in the HTN group. These results are in line with the data from previous studies on the associated risk factors of hypertension [17].

Furthermore, multivariate analysis in this study revealed that WHtR, unlike BMI and WHR, was found to be significantly associated with increased risk of hypertension, subsequent to adjustment for age, gender, history of CAD, SBP, FBS, and TG. Moreover, The ROC analysis of anthropometric indices for prediction of hypertension demonstrated that all indices, especially WHtR, exhibited a statistically significant predictive power in detecting the incidence of HTN in patients with T2DM. Therefore, according to our data, WHtR is a better screening tool for HTN than WHR and BMI in Iranian men with type 2 diabetes. This finding is similar to the results of various studies on different ethnicities worldwide [7, 18-23]. Three meta-analyses also supported WHtR as the most accurate anthropometric index for predicting hypertension [12, 24, 25]. WHtR can be considered as the more accurate obesity index. Height can predict hypertension and diabetes [26, 27], and the percentage of body fat is an independent risk factor for CVD [28]. People with shorter stature noticeably have higher amounts of body fat than taller individuals with the same BMI [29]. Individuals with similar WCs might not have a similar percentage of body fat if they are unequal in height. Therefore, WHtR can be a helpful predictor of hypertension by considering the impact of waist circumference and height on body fat composition [22]. In the current study, optimal cutoff for WHtR was calculated as 0.59. A recent Iranian study on anthropometric indices for obesity reported cutoff of 0.49-0.51 among adults [30]. Hsieh et al. recommended a WHtR cutoff of 0.5 for Japanese men and women [31]. Tseng et al. also reported a cutoff of 0.48–0.52 in both sexes and suggested that this is the clinical importance of WHtR as the same cutoff could be applied to different sexes and ethnicities [22]. WHtR is the least expensive and a simple measure that has a strong relationship with cardiovascular diseases, with a similar optimal cutoff at 0.5 in both sexes and various ethnic groups. Due to these features, WHtR can be of great value to public health and can be utilized as a standard screening tool for more accurate epidemiological data comparisons between studies. The cutoff value of 0.5 can be simply memorized and instantly employed to explain the risk of cardio-metabolic diseases to the patients to determine if the WC is less than half of the height. Thus, no more complicated calculations are needed (as required for BMI) [22].

On the other hand, Cuban and Japanese studies suggested BMI as the best single indicator for the risk of developing hypertension [9, 10]. Li et al. reported BMI and WC can predict incident hypertension more accurately than WHR, skinfold thickness, and WHtR in the Chinese population [32]. A study on the Iranian population also reported that BMI is the best predictor of hypertension in men; BMI combined with WHR were the best predictors of hypertension in women [33]. Another Iranian cross-sectional study also suggested WHR combined with BMI as the best predictor of hypertension in women [11]. Rezende et al. suggested that both central obesity (WC and WHtR) and overall obesity (BMI) anthropometric indicators could be used in the Brazilian population to evaluate the risk of incident hypertension [34]. Another Brazilian study, as well as a 2008 meta-analysis also reported that all indices had similar performances in hypertension detection [13, 35]. These differences might be due to the populations’ varying characteristics, differences in sampling strategies, data collection quality, and the differences in operational definitions for abdominal and general obesity [36]. A screening measure should be both efficient and practical. BMI needs measuring both height and weight, whereas WHtR needs WC and height. It seems that self-assessment of height is more accurate than weight [25]. As mentioned earlier, in case of a similar BMI, individuals with shorter stature have a markedly higher percentage of body fat than taller individuals [29]. In addition, some disadvantages have been reported for WC and WHR. These obesity anthropometric indices cannot reflect body height [37, 38]. Hsieh et al. argued that among the Japanese men in the third quartile of WC (84.5 -89 cm), shorter individuals were more prone to hypertension than the taller individuals [39]. Furthermore, it is harder to measure hip circumference than WC, since there is a need to accurately identify the point of maximal protrusion of the buttocks in obese people [40]. A former study argued that the area under the ROC for WHR for hypertension was reported as the lowest among anthropometric indices [41]. This finding is in line with the results of our study. Most importantly, WHR and WC are not preferable due to the variously reported cutoffs for the two sexes [37, 42].

The current study had several strengths. Firstly, to the best of our knowledge, this is the first prospective cohort study comparing obesity anthropometric indices in patients with diabetes. Secondly, it used a prospective cohort design as well as a large population-based sample. Thus, there is a clear causality between incident hypertension and obesity anthropometric indices. Thirdly, professional interviewers performed several interviews. Anthropometric data were recorded through repeated measurements in accordance with a standard protocol. Such procedures helped to reduce bias. This study had several limitations. Firstly, since our study population was limited to the Iranian ethnicity, caution should be taken when extrapolating the results to other ethnicities. Secondly, since the participants’ diet intake details were not available, we couldn’t adjust for hypertension-related covariates such as salt and fat intake. Thirdly, repeated measures of the anthropometric indices and other variables measured at baseline, were not available for all the patients; so we could not analyze the role of time-varying indices and adjust for the time-varying confounders.

## Conclusion

The present prospective study showed that WHtR is a more accurate screening tool for HTN than WHR and BMI in patients with type 2 diabetes. Our data reinforce the significance of using the most accurate anthropometric indices as a component of public health programs strategies to prevent and control the obesity epidemic and warn about the risk of developing hypertension. Therefore, WHtR could be recommended as a useful and accurate screening tool for hypertension prediction due to its high discriminative power.

## Data Availability

The data could be provided by request.

## References

1. Gill, G., et al., Intensified treatment of type 2 diabetes—positive effects on blood pressure, but not glycaemic control. Qjm, 2003. 96(11): p. 833–836.

2. Stanciu, I., et al., Clinical trial evidence for cardiovascular risk reduction in type 2 diabetes. Journal of Cardiovascular Nursing, 2002. 16(2): p. 24–43.

3. Hu, G., et al., Physical activity, cardiovascular risk factors, and mortality among Finnish adults with diabetes. Diabetes care, 2005. 28(4): p. 799–805.

4. White, F., L. Wang, and H.F. Jelinek, Management of hypertension in patients with diabetes mellitus. Experimental & Clinical Cardiology, 2010. 15(1): p. 5.

5. Chobanian, A.V., et al., Seventh report of the joint national committee on prevention, detection, evaluation, and treatment of high blood pressure. hypertension, 2003. 42(6): p. 1206–1252.

6. Mendis, S., et al., Global atlas on cardiovascular disease prevention and control. 2011: World Health Organization.

7. Caminha, T.C., et al., Waist-to-height ratio is the best anthropometric predictor of hypertension: a population-based study with women from a state of northeast of Brazil. Medicine, 2017. 96(2).

8. Liu, K., et al., Voxel-based morphometry reveals regional reductions of gray matter volume in school-aged children with short-term type 1 diabetes mellitus. Neuroreport, 2019. 30(7): p. 516–521.

9. Barbosa, A.R., et al., Anthropometric indexes of obesity and hypertension in elderly from Cuba and Barbados. The journal of nutrition, health & aging, 2011. 15(1): p. 17–21.

10. Sakurai, M., et al., Gender differences in the association between anthropometric indices of obesity and blood pressure in Japanese. Hypertension Research, 2006. 29(2): p. 75–80.

11. Faramarzi, E., et al., Determination of the Best Anthropometric Index of Obesity for Prediction of Prehypertension and Hypertension in a Large Population - Based - Study; the Azar-Cohort. Iran Red Crescent Med J, 2018. 20(3): p. e59911.

12. De Simone, G. and M. Chinali, Is central obesity a better discriminator of the risk of hypertension than body mass index in ethnically diverse populations? Commentary. Journal of hypertension, 2008. 26(2): p. 169–181.

13. Lee, C.M.Y., et al., Indices of abdominal obesity are better discriminators of cardiovascular risk factors than BMI: a meta-analysis. Journal of clinical epidemiology, 2008. 61(7): p. 646–653.

14. Unger, T., et al., 2020 International Society of Hypertension Global Hypertension Practice Guidelines. Hypertension, 2020. 75(6): p. 1334–1357.

15. Esteghamati, A., et al., Diabetes in Iran: prospective analysis from first nationwide diabetes report of National Program for Prevention and Control of Diabetes (NPPCD-2016). Scientific reports, 2017. 7(1): p. 1–10.

16. Govindarajan, G., J.R. Sowers, and C.S. Stump, Hypertension and diabetes mellitus. Journal- Hypertension and Diabetes Mellitus, 2006.

17. Singh, S., R. Shankar, and G.P. Singh, Prevalence and associated risk factors of hypertension: a cross-sectional study in urban Varanasi. International journal of hypertension, 2017. 2017.

18. Kawamoto, R., et al., Usefulness of waist-to-height ratio in screening incident hypertension among Japanese community-dwelling middle-aged and elderly individuals. Clinical Hypertension, 2020. 26(1): p. 1–9.

19. Khader, Y., et al., The performance of anthropometric measures to predict diabetes mellitus and hypertension among adults in Jordan. BMC public health, 2019. 19(1): p. 1416.

20. Lee, J.-W., et al., Anthropometric indices as predictors of hypertension among men and women aged 40–69 years in the Korean population: the Korean Genome and Epidemiology Study. BMC Public Health, 2015. 15(1): p. 1–7.

21. Saeed, A.A. and N.A. Al-Hamdan, Anthropometric risk factors and predictors of hypertension among Saudi adult population–A national survey. Journal of epidemiology and global health, 2013. 3(4): p. 197–204.

22. Tseng, C.-H., et al., Optimal anthropometric factor cutoffs for hyperglycemia, hypertension and dyslipidemia for the Taiwanese population. Atherosclerosis, 2010. 210(2): p. 585–589.

23. Mansour, A.A. and M.I. Al-Jazairi, Cut-off values for anthropometric variables that confer increased risk of type 2 diabetes mellitus and hypertension in Iraq. Archives of medical research, 2007. 38(2): p. 253–258.

24. Deng, G., et al., Associations of anthropometric adiposity indexes with hypertension risk: A systematic review and meta-analysis including PURE-China. Medicine, 2018. 97(48).

25. Ashwell, M., P. Gunn, and S. Gibson, Waist□to□height ratio is a better screening tool than waist circumference and BMI for adult cardiometabolic risk factors: systematic review and meta□analysis. Obesity reviews, 2012. 13(3): p. 275–286.

26. Lara-Esqueda, A., et al., The body mass index is a less-sensitive tool for detecting cases with obesity-associated co-morbidities in short stature subjects. Int J Obes Relat Metab Disord, 2004. 28(11): p. 1443–50.

27. Fuchs, F.D., et al., Anthropometric indices and the incidence of hypertension: a comparative analysis. Obes Res, 2005. 13(9): p. 1515–7.

28. Tseng, C.H., Body composition as a risk factor for coronary artery disease in Chinese type 2 diabetic patients in Taiwan. Circ J, 2003. 67(6): p. 479–84.

29. López-Alvarenga, J.C., et al., Short stature is related to high body fat composition despite body mass index in a Mexican population. Arch Med Res, 2003. 34(2): p. 137–40.

30. Ramezankhani, A., et al., Optimum cutoff values of anthropometric indices of obesity for predicting hypertension: more than one decades of follow-up in an Iranian population. Journal of Human Hypertension, 2018. 32(12): p. 838–848.

31. Hsieh, S., H. Yoshinaga, and T. Muto, Waist-to-height ratio, a simple and practical index for assessing central fat distribution and metabolic risk in Japanese men and women. International journal of obesity, 2003. 27(5): p. 610–616.

32. Li, N., et al., Is waist-to-height ratio superior to body mass index and waist circumference in predicting the incidence of hypertension? Annals of Nutrition and Metabolism, 2019. 74(3): p. 215–223.

33. Faramarzi, E., et al., Determination of the Best Anthropometric Index of Obesity for Prediction of Prehypertension and Hypertension in a Large Population-Based-Study; the Azar-Cohort. Iranian Red Crescent Medical Journal, 2018. 20(3).

34. Rezende, A.C., et al., Is waist-to-height ratio the best predictive indicator of hypertension incidence? A cohort study. BMC Public Health, 2018. 18(1): p. 281.

35. de Souza, A.P.A., et al., Cut-off points of anthropometric markers associated with hypertension in the Brazilian population: National Health Survey, 2013. Public health nutrition, 2019. 22(12): p. 2147–2154.

36. Wai, W.S., et al., Comparison of measures of adiposity in identifying cardiovascular disease risk among Ethiopian adults. Obesity, 2012. 20(9): p. 1887–1895.

37. Li, W.-C., et al., Waist-to-height ratio, waist circumference, and body mass index as indices of cardiometabolic risk among 36,642 Taiwanese adults. European journal of nutrition, 2013. 52(1): p. 57–65.

38. Browning, L.M., S.D. Hsieh, and M. Ashwell, A systematic review of waist-to-height ratio as a screening tool for the prediction of cardiovascular disease and diabetes: 0· 5 could be a suitable global boundary value. Nutrition research reviews, 2010. 23(2): p. 247–269.

39. Hsieh, S.D. and H. Yoshinaga, Do people with similar waist circumference share similar health risks irrespective of height? The Tohoku journal of experimental medicine, 1999. 188(1): p. 55–60.

40. Organization, W.H., Waist circumference and waist-hip ratio: report of a WHO expert consultation, Geneva, 8-11 December 2008. 2011.

41. Gupta, S. and S. Kapoor, Optimal cut-off values of anthropometric markers to predict hypertension in North Indian population. Journal of community health, 2012. 37(2): p. 441–447.

42. Ashwell, M. and S.D. Hsieh, Six reasons why the waist-to-height ratio is a rapid and effective global indicator for health risks of obesity and how its use could simplify the international public health message on obesity. International journal of food sciences and nutrition, 2005. 56(5): p. 303–307.

